# Safety of COVID-19 vaccines in pregnancy: a Canadian National Vaccine Safety (CANVAS) Network study

**DOI:** 10.1101/2022.02.22.22271358

**Authors:** Manish Sadarangani, Phyumar Soe, Hennady Shulha, Louis Valiquette, Otto G Vanderkooi, James D Kellner, Matthew P Muller, Karina A Top, Jennifer E Isenor, Allison McGeer, Mike Irvine, Gaston De Serres, Kimberly Marty, Julie A. Bettinger, Canadian Immunization Research Network

**Affiliations:** Vaccine Evaluation Center, BC Children’s Hospital Research Institute, Vancouver, British Columbia, Canada; Department of Pediatrics, University of British Columbia, Vancouver, British Columbia, Canada; Centre Intégré Universitaire de Santé et de Services Sociaux de l’Estrie-Centre Hospitalier Universitaire de Sherbrooke, Sherbrooke, Québec, Canada; Alberta Children’s Hospital Research Institute, Calgary, Alberta, Canada; Department of Pediatrics, University of Calgary, Calgary, Alberta, Canada; Department of Medicine, Unity Health Toronto, Toronto, Ontario, Canada; University of Toronto, Toronto, Ontario, Canada; Canadian Center for Vaccinology, IWK Health, Halifax, Nova Scotia, Canada; Department of Pediatrics, Dalhousie University, Halifax, Nova Scotia, Canada; College of Pharmacy, Dalhousie University, Halifax, Nova Scotia, Canada; Sinai Health System, Toronto, Ontario, Canada; BC Centre for Disease Control, Vancouver, British Columbia; CHU de Québec-Université Laval, Quebec, Québec, Canada; Institut national de santé publique du Québec, Québec, Canada

**Author notes:** **Corresponding author** Dr. Manish Sadarangani, Vaccine Evaluation Center, BC Children’s Hospital Research Institute, 950 West 28th Avenue, Vancouver BC V5Z 4H4, Canada., Ph.

## Abstract

**Background:** Pregnant individuals have been receiving COVID-19 vaccines following pre-authorization clinical trials in non-pregnant people. This study aimed to determine significant health events amongst pregnant females after COVID-19 vaccination, compared with unvaccinated pregnant controls and vaccinated non-pregnant individuals.

**Methods:** Study participants were pregnant and non-pregnant females aged 15-49 years who had received any COVID-19 vaccine, and pregnant unvaccinated controls. Participants reported significant health events occurring within seven days of vaccination. We employed multivariable logistic regression to examine significant health events associated with mRNA vaccines.

**Findings:** Overall 226/5,597(4.0%) vaccinated pregnant females reported a significant health event after dose one of an mRNA vaccine, and 227/3,108(7.3%) after dose two, compared with 11/339(3.2%) pregnant unvaccinated females. Pregnant vaccinated females had an increased odds of a significant health event after dose two of mRNA-1273 (aOR 4.4,95%CI 2.4-8.3) compared to pregnant unvaccinated controls, but not after dose one of mRNA-1273 or any dose of BNT162b2. Pregnant females had decreased odds of a significant health event compared to non-pregnant females after both dose one (aOR 0.63,95%CI 0.55-0.72) and dose two (aOR 0.62,95%CI 0.54-0.71) of mRNA vaccination. There were no significant differences in any analyses when restricted to events which led to medical attention.

**Interpretation:** COVID-19 mRNA vaccines have a good safety profile in pregnancy. Rates of significant health events were higher after dose two of mRNA-1273 compared with unvaccinated controls, with no difference when considering events leading to medical consultation. Rates of significant health events were lower in pregnant females than similarly aged non-pregnant individuals.

**Funding:** This work was supported by the COVID-19 Vaccine Readiness funding from the Canadian Institutes of Health Research and the Public Health Agency of Canada CANVAS grant number CVV-450980 and by funding from the Public Health Agency of Canada, through the Vaccine Surveillance Reference Group and the COVID-19 Immunity Task Force.

## Background

The coronavirus disease 2019 (COVID-19) pandemic has disproportionately affected pregnant people, who are at higher risk of severe disease compared with similarly aged non-pregnant individuals. Pregnancy leads to an increased risk of COVID-19 related hospitalization, intensive care unit admission, need for mechanical ventilation and death – due to changes in cardiovascular, pulmonary and immunologic physiology^1^. In addition, severe acute respiratory syndrome coronavirus-2 (SARS-CoV-2) infection results in increased risk of adverse pregnancy outcomes such as hypertension, pre-eclampsia, impaired fetal growth, and preterm birth^1^.

COVID-19 vaccines have been available in Canada since December 2020, following evidence of high efficacy in pre-authorization clinical trials^2^. Multiple expert groups published positive recommendations for use of COVID-19 vaccines in pregnancy early on in vaccine deployment, based largely on established prior safety of inactivated vaccines in pregnancy, several decades of use of vaccines in pregnancy (such as tetanus toxoid, influenza and tetanus-diphtheria-pertussis [DTP]), and reassuring data from the small number of incidental pregnancies occurring during pre-authorization trials^3,4^. Data from large cohorts requires post-implementation studies, and data has been accruing on the safety of mRNA vaccines during pregnancy^5-7^. These are primarily descriptive studies, relying on comparisons with historic rates of adverse events and without a contemporaneous control group to enable comparison with background rates of adverse events following immunization (AEFIs).

The Canadian National Vaccine Safety (CANVAS) Network was established during the 2009 influenza pandemic to provide rapid, real-time safety data during rollout of immunization programs, and has monitored the safety of seasonal influenza vaccines in Canada since then, as well as being utilized for other emergency campaigns, such as for a capsular group B meningococcal vaccine program^8^. The CANVAS Network has been monitoring COVID-19 vaccine safety in Canada since the start of the vaccine rollout, and recruitment remains ongoing^9^. Distinguishing components of the CANVAS approach are active follow-up of individuals with significant health events and active enrolment of a control group to enable comparisons with unvaccinated individuals in a similar time frame. The aim of this analysis was to compare rates of health events in (1) vaccinated pregnant females and vaccinated non-pregnant females of the same age and (2) vaccinated and unvaccinated (control) pregnant females.

## Methods

### Study population

In this ongoing study, we actively recruit participants from seven Canadian provinces and territories accounting for over 75% of the national population including Ontario, Quebec, British Columbia, Alberta, Nova Scotia, Yukon, and Prince Edward Island^9^. Vaccinated individuals can enroll in the study if: they have received the first dose of an authorized COVID-19 vaccine within the prior seven days; have an active email address and telephone number; can communicate in English or French; reside in the above-mentioned provinces/territories. Individuals can participate as controls if they are unvaccinated and fulfil the other criteria. People can contribute to both vaccinated and control groups, if they initially enroll prior to vaccination and subsequently enroll after vaccination. Individuals who enroll as control participants and are expected to be vaccinated within the next six months complete a retrospective control survey. All other control participants are followed prospectively.

### Data collection

A detailed description of recruitment procedures has been published^9^. Vaccinated participants are surveyed via email for the occurrence of AEFIs during the seven days following each dose of COVID-19 vaccine and at seven months after their first vaccine dose. All participants are asked about injection site reactions, but only those that indicate they have a significant health event are requested to provide further details. Control participants are requested to note the occurrence of health problems in the prior seven days, 28 days, and six months. We undertake telephone follow up for those who report a medically attended event on any survey. We send up to two automatic reminders every 72 hours for non-responders in both the vaccinated and unvaccinated groups. Survey data are collected in a secure REDCap (Research Electronic Data Capture) database^10,11^. Data collected include age group, sex, gender, ethnicity, self-reported prior lab-confirmed SARS-CoV-2 infection, COVID-19 disease severity using a standardized scale, pregnancy and lactation status, presence of auto-immune or immunocompromising medical conditions, general health status, COVID-19 vaccine product administered and lot number, occurrence or worsening of health events, and requirement for medical attention, and/or hospitalization related to the health event.

### Analytic samples and study variables

For the purposes of this analysis, we included all females reporting a pregnancy on any survey, and non-pregnant females in the same age groups (15-49 years), as of 4^th^ November 2021 (Figure 1). Specifically, we included vaccinated females who completed surveys after the first and/or second COVID-19 vaccine dose, and control data from individuals who completed 7-day surveys. When comparing health event rates amongst pregnant people, the same pregnant control group was used to compare with vaccinated pregnant people who received first and/or second dose of COVID-19 vaccines. For statistical modelling, the analysis was restricted to those who received any mRNA vaccines (BNT162b2 and mRNA-1273) since very few pregnant people reported receiving the ChAdox1-S vaccine for either dose one or dose two (Figure 1). For pregnant participants who reported experiencing a miscarriage following both dose one and dose two, only the first report was included to avoid duplicate counting from individuals reporting the same miscarriage twice, given the relatively short interval between vaccine doses and low likelihood of two miscarriages occurring in this time frame.

**Figure 1.**
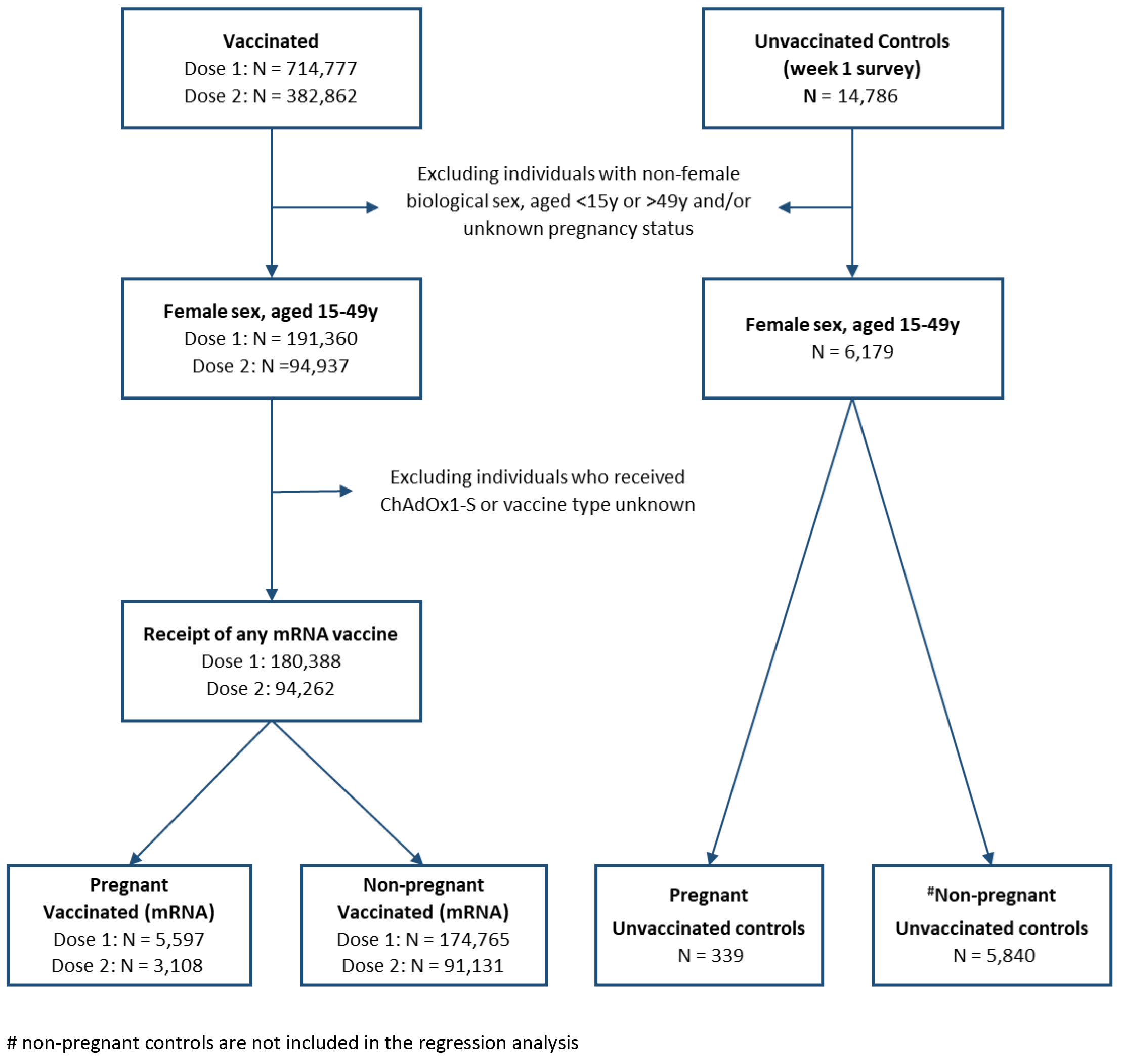
Flow chart depicting the analytical samples (dose one and dose two samples) for examining AEFIs associated with COVID-19 vaccine using data from CANVAS-COVID-19 study.

The primary endpoint was ‘significant health event’, defined as a new or worsening of a health event sufficient to cause work/school absenteeism, medical consultation and/or prevent daily activities in the previous seven days. ‘Serious health event’ was a secondary endpoint, and defined as any event resulting in emergency department visit and/or hospitalization in the previous seven days. Two types of exposures were analyzed: 1) vaccination status among pregnant people, and 2) pregnancy status among vaccinated people.

### Statistical analysis

We estimated rates of significant and serious health events including common and uncommon symptoms following the first and second doses of COVID-19 vaccines. To examine associations between the outcomes and the exposures, two sets of univariate/multivariate (MV) logistic regression models were built for each type of exposure (vaccination status among pregnant people and pregnancy status among vaccinated people). When fitting MV models, we adjusted known or expected covariates such as age group, previous COVID-19 infection, and trimester of pregnancy, as appropriate. Three different vaccine groups were evaluated: 1) BNT162b2, 2) mRNA-1273, and 3) any mRNA vaccine. The estimated odds ratio and 95% confidence intervals were reported. We conducted complete case analysis as no variable had ≥2% of missing data. We verified the absence of multicollinearity with variance inflation factor using a cut-off point of <5.

To evaluate the robustness of findings, we conducted two sensitivity analyses. In the first, our primary end point (significant health event) was restricted to new or worsening health events also resulting in medical consultation in the previous seven days. We repeated the same analysis after the first and second doses of COVID-19 vaccines. Secondly, we restricted the dataset of pregnant people to those who reported being in excellent health and compared this with the results from the primary analysis. Data cleaning was done in SAS version 9.4 (SAS Institute Inc.) and analysis was completed in R software version 4.1.1 (R foundation for Statistical Computing, Vienna, Austria).

### Ethics

All participants provided informed consent electronically. Each study site has Research Ethics Board approvals for the project (UBC Children’s & Women’s, CIUSSS de l’Estrie – CHUS, Health PEI, Conjoint Health Research Ethics Board, University of Calgary and Alberta Health Services, IWK Health, Unity Health Toronto, and CHU de Québec-Université Laval).

## Results

### Overall study population

As of 4^th^ November 2021, a total of 191,360 and 94,937 females aged 15-49 years with known pregnancy status had completed the first and second dose surveys, respectively (Figure 1). A total of 5,625 (2.9%) and 3,114 (3.3%) females reported being pregnant at the time of receiving first and second doses of COVID-19 vaccines, respectively (Supplementary Table 1). Of these, most pregnant participants had received BNT162b2 (3,414 after dose one and 1,892 after dose two) or mRNA-1273 (2,183 after dose one and 1,216 after dose two) COVID-19 vaccines and few individuals reported receiving ChAdOx1-S (25 after dose one and six after dose two). Among the enrolled unvaccinated control participants, a total of 6,179 females aged 15-49 years had completed 7-day surveys. Of these, 339 (5.5%) reported being pregnant during the survey period (Supplementary Table 1). Amongst both vaccinated and control groups, pregnant individuals in all three trimesters were well represented (Table 1).

**Table 1.** Baseline demographics characteristics with health event rates among study populations.

|  | Pregnant Control<br>N(%) | Dose 1 |  |  |  |  |  | Dose 2 |  |  |  |  |  |
| --- | --- | --- | --- | --- | --- | --- | --- | --- | --- | --- | --- | --- | --- |
|  |  | Any mRNA |  | BNT162b2 |  | mRNA-1273 |  | Any mRNA |  | BNT162b2 |  | mRNA-1273 |  |
|  |  | Preg(-)<br>N (%) | Preg(+)<br>N(%) | Preg(-)<br>N (%) | Preg(+)<br>N (%) | Preg(-)<br>N (%) | Preg(+)<br>N(%) | Preg(-)<br>N (%) | Preg(+)<br>N(%) | Preg(-)<br>N (%) | Preg(+)<br>N(%) | Preg(-)<br>N (%) | Preg(+)<br>N(%) |
| **Sample** | **339** | **174765** | **5597** | **107121** | **3414** | **67644** | **2183** | **91131** | **3108** | **53077** | **1892** | **38054** | **1216** |
| **Significant HE** | 11(3.2) | 10950(6.3) | 226(4.0) | 6501(6.1) | 137(4.0) | 4449(6.6) | 89(4.1) | 10254(11.3) | 227(7.3) | 4121(7.8) | 80(4.2) | 6133(16.1) | 147(12.1) |
| **Severe HE** | 4(1.2) | 2439(1.4) | 78(1.4) | 1392(1.3) | 45(1.3) | 1047(1.5) | 33(1.5) | 1257(1.4) | 47(1.5) | 663(1.2) | 22(1.2) | 594(1.6) | 25(2.1) |
| **Serious HE** | 2(0.6) | 733(0.4) | 31(0.6) | 421(0.4) | 18(0.5) | 312(0.5) | 13(0.6) | 343(0.4) | 19(0.6) | 192(0.4) | 8(0.4) | 151(0.4) | 11(0.9) |
| **Trimester** | 11(3.2) | 10950(6.3) | 226(4.0) | 6501(6.1) | 137(4.0) | 4449(6.6) | 89(4.1) | 10254(11.3) | 227(7.3) | 4121(7.8) | 80(4.2) | 6133(16.1) | 147(12.1) |
| 1<sup>st</sup> trimester | 5(1.5) |  | 70(1.3) |  | 42(1.2) |  | 28(1.3) |  | 52(1.7) |  | 18(1.0) |  | 34(2.8) |
| 2<sup>nd</sup> trimester | 3(0.9) |  | 96(1.7) |  | 56(1.6) |  | 40(1.8) |  | 85(2.7) |  | 29(1.5) |  | 56(4.6) |
| 3<sup>rd</sup> trimester | 2(0.6) |  | 47(0.8) |  | 34(1.0) |  | 13(0.6) |  | 74(2.4) |  | 28(1.5) |  | 46(3.8) |
| **Age groups** |  |  |  |  |  |  |  |  |  |  |  |  |  |
| 15-29 | 51(15.0) | 59263(33.9) | 1417(25.3) | 41221(38.5) | 796(23.3) | 18042(26.7) | 621(28.4) | 26324(28.9) | 716(23.0) | 17008(32.0) | 422(22.3) | 9316(24.5) | 294(24.2) |
| 30-49 | 288(85.0) | 115502(66.1) | 4180(74.7) | 65900(61.5) | 2618(76.7) | 49602(73.3) | 1562(71.6) | 64807(71.1) | 2392(77.0) | 36069(68.0) | 1470(77.7) | 28738(75.5) | 922(75.8) |
| **Ethnicity** |  |  |  |  |  |  |  |  |  |  |  |  |  |
| White | 22(6.5) | 47539(27.2) | 1818(32.5) | 29421(27.5) | 1166(34.2) | 18118(26.8) | 652(29.9) | 50779(55.7) | 1778(57.2) | 25645(48.3) | 957(50.6) | 25134(66.0) | 821(67.5) |
| Black | 2(0.6) | 1155(0.7) | 18(0.3) | 802(0.7) | 12(0.4) | 353(0.5) | 6(0.3) | 1169(1.3) | 22(0.7) | 692(1.3) | 14(0.7) | 477(1.3) | 8(0.7) |
| East Asian | 0 | 3367(1.9) | 117(2.1) | 2427(2.3) | 87(2.5) | 940(1.4) | 30(1.4) | 3471(3.8) | 106(3.4) | 2213(4.2) | 72(3.8) | 1258(3.3) | 34(2.8) |
| South Asian | 0 | 1977(1.1) | 95(1.7) | 1502(1.4) | 81(2.4) | 475(0.7) | 14(0.6) | 2041(2.2) | 104(3.3) | 1325(2.5) | 83(4.4) | 716(1.9) | 21(1.7) |
| South-East Asian | 1(0.3) | 1552(0.9) | 47(0.8) | 1121(1.0) | 36(1.1) | 431(0.6) | 11(0.5) | 1587(1.7) | 45(1.4) | 995(1.9) | 27(1.4) | 592(1.6) | 18(1.5) |
| Indigenous | 0 | 699(0.4) | 22(0.4) | 476(0.4) | 18(0.5) | 223(0.3) | 4(0.2) | 703(0.8) | 21(0.7) | 436(0.8) | 13(0.7) | 267(0.7) | 8(0.7) |
| Middle-Eastern | 0 | 1352(0.8) | 44(0.8) | 821(0.8) | 30(0.9) | 531(0.8) | 14(0.6) | 1361(1.5) | 44(1.4) | 751(1.4) | 25(1.3) | 610(1.6) | 19(1.6) |
| Latino | 0 | 1392(0.8) | 48(0.9) | 807(0.8) | 23(0.7) | 585(0.9) | 25(1.1) | 1460(1.6) | 50(1.6) | 736(1.4) | 19(1.0) | 724(1.9) | 31(2.5) |
| Mixed | 0 | 2714(1.6) | 92(1.6) | 1888(1.8) | 68(2.0) | 826(1.2) | 24(1.1) | 2800(3.1) | 88(2.8) | 1619(3.1) | 53(2.8) | 1181(3.1) | 35(2.9) |
| Unknown/Other | 314(92.6) | 113016(64.7) | 3296(58.9) | 67855(63.3) | 1893(55.4) | 45161(66.8) | 1403(64.3) | 25758(28.3) | 850(27.3) | 18664(35.2) | 629(33.2) | 7094(18.6) | 221(18.2) |
| **Health status** |  |  |  |  |  |  |  |  |  |  |  |  |  |
| Ex/VG/Good | 333(98.2) | 151446(86.7) | 5029(89.9) | 91266(85.2) | 2961(86.7) | 60180(89.0) | 2068(94.7) | 77911(85.5) | 2747(88.4) | 44362(83.6) | 1617(85.5) | 33549(88.2) | 1130(92.9) |
| Fair/poor/unk | 6(1.8) | 23319(13.3) | 568(10.1) | 15855(14.8) | 453(13.3) | 7464(11.0) | 115(5.3) | 13220(14.5) | 361(11.6) | 8715(16.4) | 275(14.5) | 4505(11.8) | 86(7.1) |
| **Province** |  |  |  |  |  |  |  |  |  |  |  |  |  |
| AB | 37(10.9) | 28987(16.6) | 778(13.9) | 22129(20.7) | 650(19.0) | 6858(10.1) | 128(5.9) | 17260(18.9) | 544(17.5) | 12052(22.7) | 422(22.3) | 5208(13.7) | 122(10.0) |
| ON | 178(52.5) | 44184(25.3) | 2080(37.2) | 36571(34.1) | 1636(47.9) | 7613(11.3) | 444(20.3) | 32324(35.5) | 1318(42.4) | 20445(38.5) | 888(46.9) | 11879(31.2) | 430(35.4) |
| QC | 4(1.2) | 65501(37.5) | 1464(26.2) | 21446(20.0) | 174(5.1) | 44055(65.1) | 1290(59.1) | 27621(30.3) | 664(21.4) | 11609(21.9) | 188(9.9) | 16012(42.1) | 476(39.1) |
| BC/YK | 85(25.1) | 31287(17.9) | 1132(20.2) | 23197(21.7) | 834(24.4) | 8090(11.9) | 298(13.6) | 10393(11.4) | 484(15.6) | 7437(14.0) | 341(18.0) | 2956(7.8) | 143(11.8) |
| PEI/NS | 35(10.3) | 4806(2.7) | 143(2.5) | 3778(3.5) | 120(3.6) | 1028(1.5) | 23(1.1) | 3533(3.8) | 98(3.1) | 1534(2.9) | 53(2.8) | 1999(5.2) | 45(3.7) |
| **History of COVID-19 infection** |  |  |  |  |  |  |  |  |  |  |  |  |  |
| No | 331(97.6) | 167100(95.8) | 5434(97.2) | 102013(95.4) | 3310(97.1) | 65087(96.4) | 2124(97.3) | 87891(96.5) | 3009(96.9) | 50993(96.1) | 1821(96.3) | 36898(97.0) | 1188(97.7) |
| Yes | 8(2.4) | 7416(4.2) | 158(2.8) | 4952(4.6) | 99(2.9) | 2464(3.6) | 59(2.7) | 3181(3.5) | 97(3.1) | 2055(3.9) | 69(3.7) | 1126(3.0) | 28(2.3) |

HE: Health event; Ex: Excellent; VG: Very Good
Significant HE: a new or worsening of a health event sufficient to cause work/school absenteeism, medical consultation and/or prevent daily activities in the previous seven days
Severe HE: a subgroup of significant HEs – a new or worsening of a health event resulting in medical consultation in the previous seven days
Serious HE: any event resulting in emergency department visit and/or hospitalization in the previous seven days
AB: Alberta, ON: Ontario, QC: Quebec, BC/YK: British Columbia/Yukon, PEI/NS: Prince Edward Island/Nova Scotia

Most respondents in both control and vaccinated groups were aged 30-49 (62-85% across the various groups) and reported being in excellent/very good/good health (85-98%) (Table 1). The ethnic origin of more than half of participants in both vaccinated and control groups was unknown because the question regarding ethnicity was introduced only in the dose two survey questionnaires and was optional. Among those who reported their ethnicity, the majority were White, followed by East Asian and South Asian. Prevalence of previous COVID-19 infection varied between 2.3% and 4.6% (Table 1).

### Health events in pregnant people: vaccinated pregnant *vs*. pregnant unvaccinated controls

Overall, 226 (4.0%) mRNA-vaccinated pregnant females reported a significant health event within seven days after dose one of an mRNA vaccine, and 227 (7.3%) after dose two (Table 1). The rates were similar after dose one for both mRNA vaccines (4.0% for BNT162b2 and 4.1% for mRNA-1273), but higher for mRNA-1273 (12.1%) than BNT162b2 (4.2%) after dose two. The most common significant health events after dose two of mRNA-1273 in pregnant females were feeling unwell/malaise/myalgia (n=139; 11.4%), headache/migraine (n=103; 8.5%) and respiratory tract infection (n=68; 5.6%) (Table 2). Among pregnant vaccinees who reported experiencing significant health events, most of them recognized their symptoms within 24 hours following vaccination (75-79% after dose one and 86-95% after dose two of an mRNA vaccine) and the majority (69-85%) resolved within three days (Supplementary Table 2). In comparison, 11 (3.2%) pregnant unvaccinated participants (controls) reported similar events in the seven days prior to survey completion (Table 1). Of these, 18.2% reported their symptoms 24 hours prior to the control survey and 82% of them reported their events as on-going for at least six days (Supplementary Table 2). The reported serious health events were rare (0.6-0.9% across various pregnant groups; Table 1) and occurred at similar rates in vaccinated pregnant individuals and unvaccinated controls after dose one and dose two for all vaccine types (Table 1, Supplementary Table 3).

**Table 2.** Injection site reactions and details of significant health events reported in the seven days following COVID-19 vaccination in vaccinated pregnant females and unvaccinated controls.

|  | Dose 1 |  |  |  | Dose 2 |  |  |
| --- | --- | --- | --- | --- | --- | --- | --- |
|  | Control<br>N (%) | Any mRNA<br>N (%) | BNT162b2<br>N (%) | mRNA-1273<br>N (%) | Any mRNA<br>N (%) | BNT162b2<br>N (%) | mRNA-1273<br>N (%) |
| **Total N** | **339** | **5597** | **3414** | **2183** | **3108** | **1892** | **1216** |
| **Injection Site Reactions\*** | **N/A** | **4515(80.7)** | **2631(77.2)** | **1884(86.3)** | **2440(78.5)** | **1430(75.6)** | **1010(83.1)** |
| **Any significant health events\*** | **11(3.2)** | **226(4.0)** | **137(4.0)** | **89(4.1)** | **227(7.3)** | **80(4.2)** | **147(12.1)** |
| **General** |  |  |  |  |  |  |  |
| Feeling unwell/malaise/myalgia | 3(0.9) | 168(2.9) | 104(3.0) | 64(2.9) | 205(6.6) | 66(3.5) | 139(11.4) |
| Fever | 1(0.3) | 34(0.6) | 21(0.6) | 13(0.6) | 73(2.3) | 18(1.0) | 55(4.5) |
| Arthritis/joint pain | 2(0.6) | 31(0.6) | 16(0.5) | 15(0.7) | 57(1.8) | 19(1.0) | 38(3.1) |
| Jaundice | 0 | 1(0.0) | 0 | 1(0.0) | 0 | 0 | 0 |
| **Neurologic** |  |  |  |  |  |  |  |
| Headache/migraine | 2(0.6) | 128(2.3) | 82(2.4) | 46(2.1) | 144(4.6) | 41(2.2) | 103(8.5) |
| Dizziness/vertigo | 1(0.3) | 49(0.9) | 30(0.9) | 19(0.9) | 41(1.3) | 16(0.8) | 25(2.1) |
| Paresthesia | 0 | 24(0.4) | 16(0.5) | 8(0.4) | 27(0.9) | 12(0.6) | 15(1.2) |
| Fainting | 0 | 8(0.1) | 4(0.1) | 4(0.2) | 2(0.1) | 0 | 2(0.2) |
| Inability to walk | 0 | 7(0.1) | 2(0.1) | 5(0.2) | 8(0.3) | 2(0.1) | 6(0.5) |
| Loss of taste/smell | 0 | 10(0.2) | 9(0.3) | 1(0.0) | 4(0.1) | 1(0.1) | 3(0.2) |
| Loss of vision | 0 | 4(0.1) | 2(0.1) | 2(0.1) | 1(0.0) | 0 | 1(0.1) |
| Sudden unilateral facial weakness/paralysis | 0 | 1(0.0) | 0 | 1(0.0) | 3(0.1) | 2(0.1) | 1(0.1) |
| Seizure/convulsion | 0 | 1(0.0) | 0 | 1(0.0) | 0 | 0 | 0 |
| Other neurologic** | 0 | 9(0.2) | 5(0.1) | 4(0.2) | 3(0.1) | 1(0.1) | 2(0.2) |
| **Infection** |  |  |  |  |  |  |  |
| Respiratory tract infection | 3(0.9) | 91(1.6) | 61(1.8) | 30(1.4) | 98(3.2) | 30(1.6) | 68(5.6) |
| Shingles | 0 | 1(0.0) | 0 | 1(0.0) | 1(0.0) | 0 | 1(0.1) |
| Urinary tract infection symptoms | 0 | 6(0.1) | 2(0.1) | 4(0.2) | 2(0.1) | 0 | 2(0.2) |
| **Cardio-respiratory** |  |  |  |  |  |  |  |
| Chest pain/palpitations | 1(0.3) | 38(0.7) | 24(0.7) | 14(0.6) | 26(0.8) | 7(0.4) | 19(1.6) |
| Difficulty in breathing | 3(0.9) | 29(0.5) | 17(0.5) | 12(0.5) | 17(0.5) | 5(0.3) | 12(1.0) |
| Cough | 2(0.6) | 27(0.5) | 19(0.6) | 8(0.4) | 15(0.5) | 8(0.4) | 7(0.6) |
| Chest tightness | 0 | 20(0.4) | 13(0.4) | 7(0.3) | 17(0.5) | 6(0.3) | 11(0.9) |
| **Allergic-like** |  |  |  |  |  |  |  |
| Hives | 1(0.3) | 14(0.3) | 8 | 6(0.3) | 10(0.3) | 5(0.3) | 5(0.4) |
| Itchy eyes | 0 | 10(0.2) | 5(0.1) | 5(0.2) | 8(0.3) | 2(0.1) | 6(0.5) |
| Throat swelling | 0 | 6(0.1) | 3(0.1) | 3(0.1) | 2(0.1) | 1(0.1) | 1(0.1) |
| Redness of both eyes | 0 | 3(0.1) | 1(0.0) | 2(0.1) | 2(0.1) | 0 | 2(0.2) |
| Facial swelling | 0 | 2(0.0) | 1(0.0) | 1(0.0) | 3(0.1) | 0 | 3(0.2) |
| Eyelid swelling | 0 | 2(0.0) | 1(0.0) | 1(0.0) | 1(0.0) | 0 | 1(0.1) |
| Anaphylaxis | 0 | 1(0.0) | 0 | 1(0.0) | 0 | 0 | 0 |
| **Coagulation symptoms\*\*\*** | **0** | **6(0.1)** | **3(0.1)** | **3(0.1)** | **4(0.1)** | **1(0.1)** | **3(0.2)** |

\*All participants were asked about injection site reactions; only those who indicated a significant health event were further asked to provide details – participants are able to select multiple symptoms; \*\*Weakness or paralysis of the arms or legs/confusion/change in personality/behavior or difficulty with urination or defecation; \*\*\*Symptoms of blood clot or bleeding: swelling/pain in legs/blood clot/ bruising or pinpoint dark rash

Miscarriage/stillbirth was the most frequently reported adverse pregnancy outcome and it was reported at similar rates between control (n=7, 2.1%) and vaccinated groups within seven days after dose one of any mRNA vaccine (n=83, 1.5%) (Table 3). Almost all pregnancy losses (73/83; 88%) occurred during the first trimester. There were an additional 175/3,114 individuals who reported experiencing miscarriage or stillbirth between first COVID-19 vaccine dose and completion of the second (dose two) survey (up to 10 days after dose two), although precise timing of these events relative to vaccination was not collected. Other adverse pregnancy outcomes such as vaginal bleeding, abnormal fetal heart rate and reduce fetal movement were rarely reported within seven days following any mRNA vaccination (Table 3).

**Table 3.** Reported pregnancy outcomes in the seven days following first dose COVID-19 vaccination in vaccinated pregnant and unvaccinated pregnant controls.

| Outcome | Control<br>N (%) | Dose 1 |  |  |
| --- | --- | --- | --- | --- |
|  |  | Any mRNA<br>N (%) | BNT162b2<br>N (%) | mRNA-1273 N<br>(%) |
|  | **339** | **5,597** | **3,414** | **2,183** |
| **Miscarriage/still birth** | 7(2.1) | 81(1.5) | 51(1.5) | 32(1.5) |
| 1<sup>st</sup> trimester | 7(100.0) | 73(88.0) | 47(92.2) | 26(81.3) |
| 2<sup>nd</sup> trimester | 0 | 7(8.4) | 4(7.8) | 3(9.4) |
| 3<sup>rd</sup> trimester | 0 | 1(1.2) | 0 | 1(3.1) |
| Unknown | 0 | 2(2.4) | 0 | 2(6.3) |
| **Any adverse pregnancy outcome** | 2(0.6) | 50(0.9) | 31(0.9) | 19(0.9) |
| Preterm labour | 0 | 3(0.1) | 1(0.0) | 2(0.1) |
| High blood pressure | 0 | 6(0.1) | 4(0.1) | 2(0.1) |
| Vaginal spotting or vaginal bleeding | 1(0.3) | 21(0.4) | 10(0.3) | 11(0.5) |
| Abnormal fetal heart rate | 0 | 2(0.0) | 1(0.0) | 1(0.0) |
| Other pregnancy complications* | 1(0.3) | 30(0.5) | 17(0.5) | 13(0.6) |

Participants were able to report multiple outcomes
\*included lower abdominal pain, reduce fetal movement, cramp, vomiting etc.

In MV analysis, adjusting for age group, prior COVID-19 infection and trimester, we observed an increased risk of significant health events within seven days after the second dose of any mRNA vaccine (adjusted OR [aOR]: 2.4; 95% CI: 1.3-4.5) or the second dose of mRNA-1273 COVID-19 vaccination (aOR: 4.4, 95% CI: 2.4-8.3) amongst pregnant vaccinated individuals, compared with pregnant unvaccinated controls (Figure 2A). First dose of any mRNA vaccine (mRNA-1273 or BNT162b2) and either dose of BNT162b2 was not associated with increased risk of significant health events. Similarly, we found no significant association between vaccination status and serious health events in pregnant people (Figure 2A).

**Figure 2.**
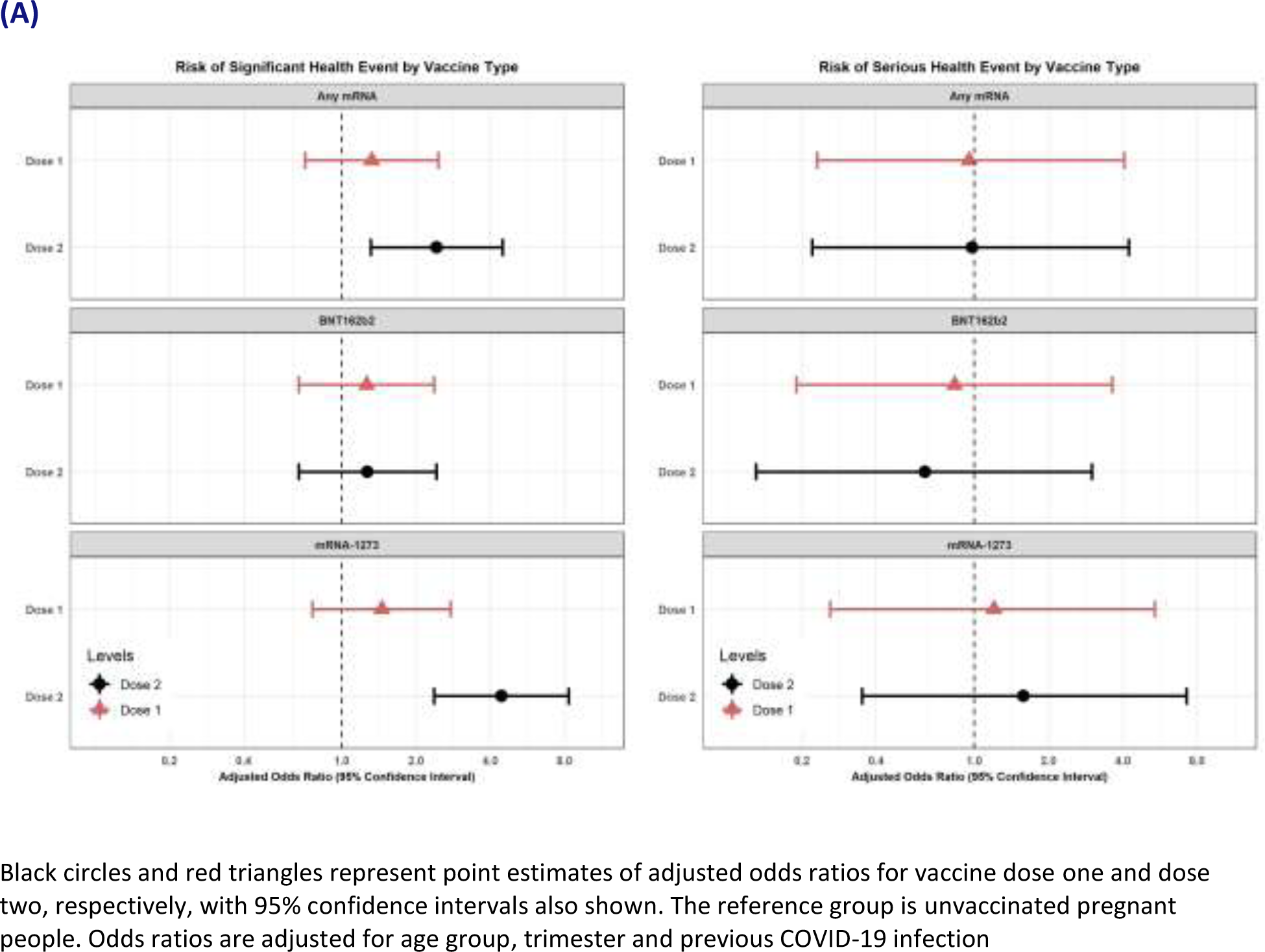
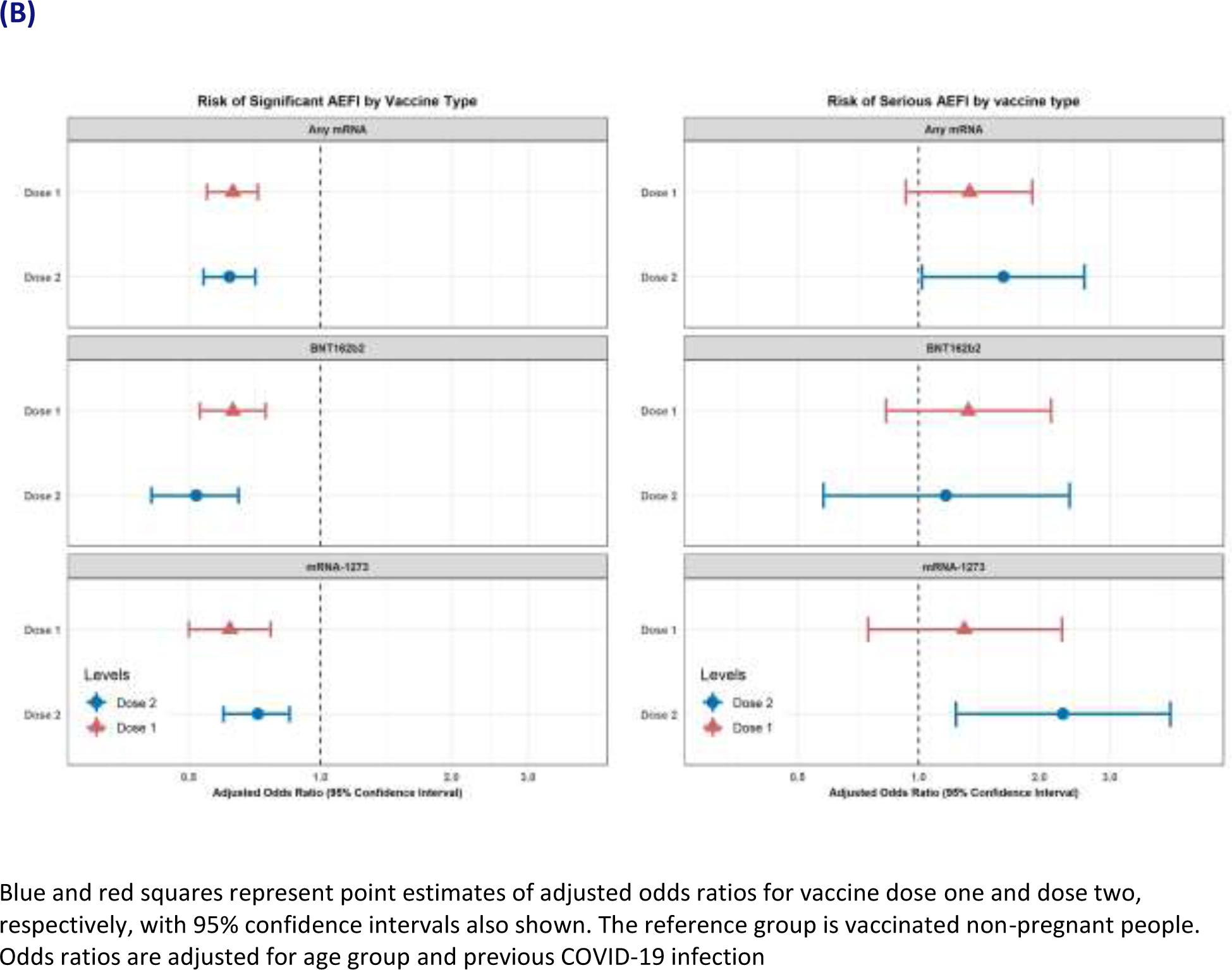
Multivariable logistic regression analyses comparing significant and serious health events amongst (A) pregnant people, comparing vaccinated with unvaccinated individuals and (B) vaccinated people, comparing pregnant with non-pregnant individuals.

In sensitivity analysis, both univariate and MV models among pregnant individuals with excellent health status revealed similar findings as in the main analysis. The increase in significant health events after dose two of any mRNA vaccine and mRNA-1273was no longer observed when we restricted our outcome variable to events requiring medical consultation – any mRNA vaccine aOR 1.26 (95% CI 0.45-3.52) and mRNA-1273 vaccine aOR 1.81 (95% CI 0.62) (Supplementary Table 4).

### Health events in vaccinated individuals: pregnant *vs*. non-pregnant

Injection site reactions such as redness, pain or swelling were reported in 76-86% of vaccine recipients. The most frequently reported severe health events within seven days of each vaccine dose were feeling unwell/malaise/myalgia (3-11%) and headache (2-9%) (Table 2). When comparing vaccinated pregnant and vaccinated non-pregnant people, significant AEFI rates (i.e., excluding injection site reactions) were consistently lower among pregnant people across all mRNA vaccine types and doses (Table 1). Overall, 4.0% and 7.3% of pregnant people reported a significant AEFI after dose one and dose two, respectively, compared with 6.3% and 11.3% for non-pregnant people. Similar differences were seen for both BNT162b2 and mRNA-1273 after both dose one and dose two. The reported rates of serious health events were low (<1%) and similar in vaccinated pregnant people compared to vaccinated non-pregnant controls (Table 1).

The MV models revealed that pregnancy was associated with a decreased risk of significant health events for any mRNA vaccine for both dose one (any mRNA aOR 0.63, 95% CI 0.55-0.72; BNT162b2 aOR 0.63, 95%CI 0.53-0.75; mRNA-1273 aOR 0.62, 95%CI 0.5-0.77) and dose two (any mRNA aOR 0.62, 95%CI 0.54-0.71; BNT162b2 aOR 0.52, 95%CI 0.41-0.65; mRNA-1273 aOR 0.72, 95%CI 0.6-0.85) (Figure 2B). However, for the secondary endpoint of serious AEFI, we found the second dose of mRNA-1273 was associated with a higher risk of serious AEFI (aOR 2.3; 95% CI: 1.2-4.2) in pregnant individuals compared with non-pregnant individuals. This association was not observed in individuals who received any mRNA or BNT162b2 for dose two or with any vaccine groups for dose one.

Sensitivity analysis that restricted our primary end point (significant health event) to those who sought medical care within seven days following each vaccine dose yielded no significant association between any mRNA vaccines and pregnancy status among vaccinated individuals (Supplementary Table 5).

## Discussion

In our prospective study collecting real-time data from pregnant individuals who were both vaccinated and unvaccinated, we found that significant health events – new or worsening health events following vaccination sufficient to cause work/school absenteeism, medical consultation and/or prevent daily activities – were lower in pregnant people than in age-matched non-pregnant vaccine recipients. Amongst pregnant individuals, significant AEFI was higher in those who received mRNA-1273 vaccine for their second dose, compared with unvaccinated pregnant people, with no difference observed for BNT162b2 after either dose. When restricted to events resulting in medical consultation, there was no difference between groups in any analyses. These data can be used to appropriately inform pregnant people regarding reactogenicity of COVID-19 vaccines during pregnancy, and should be considered alongside effectiveness and immunogenicity data to make appropriate recommendations about best use of COVID-19 vaccines in pregnancy.

The largest study to date of COVID-19 vaccine reactogenicity in pregnancy reported on approximately 30,000 pregnant people in the USA who had received BNT162b2 or mRNA-1273, using data from the v-safe registry^7^. Pregnant people in this study reported high rates of injection site pain (92% after dose two), fatigue (72%), headache (55%), myalgia (54%) and fever/chills (35-37%), with higher rates of AEFIs after dose two than dose one. The data also suggest an increased rate of these events after mRNA-1273 compared with BNT162b2 after dose two, but no formal comparisons between vaccines was done. Of note, this study collected data on all AEFI and specific information regarding severity was not reported, so it is not possible to directly compare with our study, although it is perhaps to be expected that if total AEFI rates are higher after dose two, and after mRNA-1273, then rates of significant AEFIs would follow a similar pattern. A more recent study of 7,800 pregnant people compared with 2,900 non-pregnant individuals also reported higher rates of AEFIs after dose two of COVID-19 mRNA vaccines^12^. Some AEFIs occurred at lower rates in non-pregnant individuals, including fever. Again, however, specific data on significant AEFIs were not comprehensively reported, although it was noted that 156 pregnant individuals (2.5%) sought medical care after their 2^nd^ vaccine dose, compared with only 1.5% in our cohort. A systematic review including the first study mentioned here and multiple other smaller studies reported similarly higher rates of AEFIs after dose two of COVID-19 vaccines, but no differences between pregnant and non-pregnant controls^5^.

Initial clinical trials of mRNA-1273 and BNT162b2 reported relatively high rates of AEFIs compared with most routinely administered vaccines, including higher rates for dose two than dose one^13,14^. It is therefore unsurprising that our analysis revealed similar patterns amongst pregnant individuals – although we have been able to specifically quantify the significant and serious AEFI rates in this population for each of the mRNA vaccines. Previous studies of influenza vaccines conducted using the same CANVAS methodology similarly reported higher rates of AEFIs amongst vaccinated compared with unvaccinated control participants^8^.

The lower rate of significant AEFIs amongst pregnant people, compared with vaccinated non-pregnant individuals, is important and interesting. Prior to the COVID-19 pandemic, influenza and tetanus-diphtheria-pertussis (Tdap) vaccines have been routinely recommended for pregnant people in many countries, including Canada. Previous studies have mostly reported no significant differences in AEFIs between pregnant and non-pregnant individuals^15-18^, or higher rates in pregnancy – for example, one study reported increased moderate/severe injection-site pain in pregnant (17.9%) compared with non-pregnant (11.1%) women after Tdap vaccination^16^. A number of highly dynamic immunologic adaptations occur during pregnancy, including a skew towards a Th2-dominant state^19^. Since mRNA vaccines have been specifically designed to elicit a Th1-biased immune response^20^, it is possible that the Th2-bias during pregnancy is partly responsible for this lower rate of significant AEFIs.

Our study has a number of strengths and limitations. It is a multi-centre study from across Canada and thus includes broad representation of individuals, although the majority of participants who reported ethnicity were White and these data may thus not be fully generalizable to other populations. Participants were enrolled from all trimesters in pregnancy and this was included as a co-variate in the MV models. A significant advantage over other similar studies is the contemporaneous recruitment of non-pregnant vaccinated individuals, enabling a robust direct comparison in significant AEFIs between pregnant and non-pregnant individuals. In this study we have focused on events occurring within the first seven days following vaccination and thus acute and local reactions. Longer-term follow-up of this cohort is ongoing and we will be able to comment on health events that occur on a longer time frame after vaccination once those data are available. CANVAS is based on self-report from study participants, without verification from medical records. This is subjective and may be subject to recall bias, but has been shown to be reliable for short time periods, such as used in this study^21,22^. The study relied on individuals having an email address and actively enrolling. Such individuals may differ in health seeking behavior from the rest of the population, but at least for our comparisons between groups it would be expected that they would be similarly affected. Finally, our sample size precludes detection of very rare adverse events, which rely on being identified from the general population from the passive surveillance systems in place across Canada and in other countries.

Our data provide reassuring evidence that COVID-19 mRNA vaccines are safe in pregnancy, with lower rates of significant AEFIs in pregnant people than non-pregnant vaccine recipients for both mRNA vaccines in use in Canada, after dose one and dose two. While rates of significant AEFI were highest after dose two for mRNA-1273 recipients, both mRNA vaccines are highly immunogenic and effective in pregnancy. Given the increased rate of significant complications related to COVID-19 in pregnancy, high vaccine coverage in this group is important for protection of the pregnant person and young infant, via passive transplacental transfer of antigen-specific IgG antibody and protection via breast milk. These data can, however, be used to appropriately inform pregnant individuals of the expected adverse events after vaccination. Further studies of non-COVID-19 mRNA vaccines are required to identify if the reduced reactogenicity observed in pregnant people in this study is a feature of the mRNA vaccine platform, or of these specific vaccines. Further long-term data are eagerly awaited from this cohort following 6-month follow-up. Finally, the number of pregnant individuals receiving the ChAdOx-S vaccine in our population was very low and data for this vaccine in countries where it has been more widely used are important to provide a more complete overview of the safety of COVID-19 vaccines in pregnancy.

## Supporting information

STROBE checklist

## Data Availability

De-identified data collected for the study (with data dictionary) may be made available upon approval by the study investigators, with relevant agreements (e.g., data sharing agreement) and approvals (e.g., relevant ethics approvals). Requests should be directed to the corresponding author in the first instance.

## Acknowledgements

This work was supported by the COVID-19 Vaccine Readiness funding from the Canadian Institutes of Health Research and the Public Health Agency of Canada CANVAS grant number CVV-450980 and by funding from the Public Health Agency of Canada, through the Vaccine Surveillance Reference Group and the COVID-19 Immunity Task Force. MS is supported via salary awards from the BC Children’s Hospital Foundation, the Canadian Child Health Clinician Scientist Program and the Michael Smith Foundation for Health Research.

## Author contributions statement

JB conceived and obtained funding for the study. MS, LV, OGV, JDK, MPM, KAT, JEI, AM, GDS and JB collected data and contributed to study design. KM contributed to study design. PS, HS and MI analyzed the data. MS, PS, HS, MI and JB accessed and verified the data. MS and PS drafted the original version of the manuscript. All authors had full access to the data, reviewed the manuscript, contributed to data interpretation, approved the final version and accept responsibility to submit for publication.

## Supplementary Tables

**Supplementary Table 1.** Characteristics of females aged 15-49 years in the CANVAS-COVID-19 study.

|  | Dose 1* |  |  |  |  | Dose 2* |  |  |  |
| --- | --- | --- | --- | --- | --- | --- | --- | --- | --- |
|  | Control<br>N (%) | All vaccine type<br>N (%) | BNT162b2<br>N (%) | mRNA-1273<br>N(%) | ChAdOx1-S<br>N(%) | All vaccine type<br>N(%) | BNT162b2<br>N (%) | mRNA-1273<br>N(%) | ChAdOx1-S<br>N(%) |
| **Total N** | 6,179(100) | 191,360(100) | 110,535(100) | 69,827(100) | 10,857(100) | 94,937(100) | 54,969(100) | 39,270(100) | 631(100) |
| **Non-pregnant** | 5840(94.5) | 185,735(97.1) | 107,121(96.9) | 67,644(96.9) | 10,832(99.8) | 91,823(96.7) | 53,077(96.6) | 38,054(96.9) | 625(99.2) |
| **Pregnant** | 339(5.5) | 5,625(2.9) | 3,414(3.1) | 2,183(3.1) | 25(0.2) | 3,114(3.3) | 1,892(3.4) | 1,216(3.1) | 6(0.8) |
| **Trimester** |  |  |  |  |  |  |  |  |  |
| First | 105(1.7) | 1,297(0.7) | 802(0.8) | 487(0.7) | 7 (0.1) | 956(1.0) | 568(1.0) | 385(1.0) | 3(0.3) |
| Second | 144(2.3) | 2,409(1.3) | 1,473(1.3) | 924(1.3) | 11(0.1) | 1,037(1.1) | 636(1.2) | 399(1.0) | 2(0.3) |
| Third | 90(1.5) | 1,917(1.0) | 1,139(1.0) | 770(1.1) | 7(0.1) | 1,119(1.2) | 687(1.2) | 431(1.1) | 1(0.1) |
| Unknown | 0 | 2(0.0) | 0 | 2(0.0) | 0 | 2(0.0) | 1(0.0) | 1(0.0) | 0 |

\*There were 141 females with unknown vaccine type in dose one and 67 females in dose two.

**Supplementary Table 2.** Significant and serious health event rates by vaccine type among vaccinated and unvaccinated (controls) pregnant females, and onset and duration of significant health events.

|  | Dose 1 |  |  |  | Dose 2 |  |  |
| --- | --- | --- | --- | --- | --- | --- | --- |
|  | Control<br>N (%) | Any mRNA<br>N (%) | BNT162b2<br>N (%) | mRNA-1273<br>N (%) | Any mRNA<br>N (%) | BNT162b2<br>N (%) | mRNA-1273<br>N (%) |
| **Sample size** | 339 | 5597 | 3414 | 2183 | 3108 | 1892 | 1216 |
| **Significant health events** | 11(3.2) | 226(4.0) | 137(4.0) | 89(4.1) | 227(7.3) | 80(4.2) | 147(12.1) |
| **Onset of significant health events** | 0(0.0) | 12(5.3) | 8(5.8) | 4(4.5) | 4(1.8) | 1(1.3) | 3(2.0) |
| Within 60 minutes | 2(18.2) | 118(52.2) | 69(50.4) | 49(55.1) | 183(80.6) | 55(68.8) | 128(87.1) |
| 1-24 hours | 0(0.0) | 55(24.3) | 34(24.8) | 21(23.6) | 26(11.5) | 14(17.5) | 12(8.2) |
| 2-3 days | 3(27.3) | 22(9.7) | 12(8.8) | 10(11.2) | 5(2.2) | 3(3.8) | 2(1.4) |
| 4-5 days | 4(36.4) | 11(4.9) | 9(6.6) | 2(2.2) | 4(1.8) | 4(5.0) | 0(0.0) |
| 6-7 days | 2(18.2) | 7(3.1) | 4(2.9) | 3(3.4) | 5(2.2) | 3(3.8) | 2(1.4) |
| 8+ days |  |  |  |  |  |  |  |
| **Duration of significant health events** | 0(0.0) | 6(2.7) | 4(2.9) | 2(2.2) | 2(0.9) | 1(1.3) | 1(0.7) |
| Within 60 minutes | 0(0.0) | 17(7.5) | 14(10.2) | 3(3.4) | 15(6.6) | 6(7.5) | 9(6.1) |
| 1-10 hours | 0(0.0) | 46(20.4) | 29(21.2) | 17(19.1) | 84(37.0) | 29(36.3) | 55(37.4) |
| 11-24 hours | 1(9.1) | 55(24.3) | 32(23.4) | 23(25.8) | 67(29.5) | 19(23.8) | 48(32.7) |
| 2-3 days | 1(9.1) | 33(14.6) | 17(12.4) | 16(18.0) | 18(7.9) | 6(7.5) | 12(8.2) |
| 4-5 days | 0(0.0) | 17(7.5) | 12(8.8) | 5(5.6) | 25(11.0) | 11(13.8) | 14(9.5) |
| 6+ days | 9(81.8) | 51(22.6) | 28(20.4) | 23(25.8) | 16(7.0) | 8(10.0) | 8(5.4) |
| Ongoing |  |  |  |  |  |  |  |

**Supplementary Table 3.** Reported diagnosis for serious health event rates by vaccine type among vaccinated and unvaccinated pregnant females.

|  | Dose 1 |  |  |  | Dose 2 |  |  |
| --- | --- | --- | --- | --- | --- | --- | --- |
|  | Control<br>N (%) | Any mRNA<br>N (%) | BNT162b2<br>N (%) | mRNA-1273<br>N (%) | Any mRNA<br>N (%) | BNT162b2<br>N (%) | mRNA-1273<br>N (%) |
| **Sample size** | 339 | 5597 | 3414 | 2183 | 3108 | 1892 | 1216 |
| **Serious health events** | 2(0.6) | 31(0.6) | 18(0.5) | 13(0.6) | 19(0.6) | 8(0.4) | 11(0.9) |
| **Reported diagnosis** |  |  |  |  |  |  |  |
| Anaphylaxis | 0 | 5(16.1) | 3(16.7) | 2(15.4) | 2(10.5) | 0 | 2(18.2) |
| Sepsis | 0 | 0 | 0 | 0 | 1(5.3) | 0 | 1(9.1) |
| Coagulation | 0 | 1(3.2) | 0 | 1(7.7) | 0 | 0 | 0 |
| Pulmonary Embolism | 0 | 1(3.2) | 0 | 1(7.7) | 0 | 0 | 0 |
| Bell's Palsy | 0 | 0 | 0 | 0 | 1(5.3) | 1(12.5) | 0 |
| Severe Morning sickness | 0 | 1(3.2) | 0 | 1(7.7) | 0 | 0 | 0 |
| Pre-term labor | 0 | 1(3.2) | 1(5.6) | 0 | 2(10.5) | 1(12.5) | 1(9.1) |
| Still birth | 0 | 0 | 0 | 0 | 1(5.3) | 1(12.5) | 0 |
| Miscarriage | 0 | 1(3.2) | 0 | 1(7.7) | 3(15.8) | 0 | 3(27.3) |
| Abruptio placenta | 0 | 2(6.5) | 2(11.1) | 0 | 2(10.5) | 2(25.0) | 0 |
| Fibroid pain | 0 | 2(6.5) | 2(11.1) | 0 | 0 | 0 | 0 |
| COVID-19 infection | 1(50.0) | 1(3.2) | 1(5.6) | 0 | 0 | 0 | 0 |
| Oral Thrush | 0 | 0 | 0 | 0 | 1(5.3) | 1(12.5) | 0 |
| Muscle soreness | 0 | 0 | 0 | 0 | 1(5.3) | 1(12.5) | 0 |
| Kidney stones/Cholestasis | 0 | 2(6.5) | 2(11.1) | 0 | 0 | 0 | 0 |
| Anxiety attack | 0 | 1(3.2) | 0 | 1(7.7) | 0 | 0 | 0 |
| No diagnosis | 1(50.0) | 12(38.7) | 7(38.9) | 5(38.5) | 5(26.3) | 1(12.5) | 4(36.4) |
| Prefer not to answer | 0 | 1(3.2) | 0 | 1(7.7) | 0 | 0 | 0 |

Serious health event: any event resulting in emergency department visit and/or hospitalization in the previous seven days

**Supplementary Table 4.** Results of primary and sensitivity analyses assessing the relationship between vaccination status and significant health events among pregnant people.

|  | Dose 1 |  | Dose 2 |  |
| --- | --- | --- | --- | --- |
|  | Unadjusted<br>OR (95% CI) | Adjusted<br>aOR* (95% CI) | Unadjusted<br>OR (95% CI) | Adjusted<br>aOR* (95% CI) |
| **Primary analysis<sup>a</sup>:** |  |  |  |  |
| Any mRNA | 1.25[0.68 – 2.32] | 1.32[0.71 – 2.46] | 2.35[1.27 – 4.35] | 2.42[1.31 – 4.49] |
| BNT162b2 | 1.25[0.67 – 2.33] | 1.26[0.67 – 2.37] | 1.32[0.69 – 2.50] | 1.27[0.67 – 2.42] |
| mRNA-1273 | 1.27[0.67 – 2.40] | 1.45[0.76 – 2.76] | 4.10[2.19 – 7.66] | 4.44[2.37 – 8.31] |
| **Sensitivity analysis<sup>b</sup>** |  |  |  |  |
| Any mRNA | 1.18 [0.43 -3.25] | 1.22[0.44 – 3.36] | 1.29[0.46 – 3.59] | 1.26[0.45 – 3.52] |
| BNT162b2 | 1.12[0.40 – 3.13] | 1.11[0.39 – 3.12] | 0.99[0.34 – 2.88] | 0.90[0.31 – 2.64] |
| mRNA-1273 | 1.29[0.45 – 3.65] | 1.41[0.49 – 4.07] | 1.76[0.61 – 5.09] | 1.81[0.62 – 5.28] |

<sup>a</sup>Primary end point (significant health event): a new or worsening of a health event sufficient to cause work/school absenteeism, medical consultation and/or prevent daily activities in the previous seven days
<sup>b</sup>Severe health event: a new or worsening of a health event resulting medical consultation in the previous seven days
\*Reference: unvaccinated pregnant control; adjusted for age groups, trimester and previous COVID-19 infection

**Supplementary Table 5.** Results of primary and sensitivity analyses assessing the relationship between pregnancy status and significant health events among vaccinated people.

|  | Dose 1 |  | Dose 2 |  |
| --- | --- | --- | --- | --- |
|  | Unadjusted<br>OR (95% CI) | Adjusted<br>aOR* (95% CI) | Unadjusted<br>OR (95% CI) | Adjusted<br>aOR* (95% CI) |
| **Primary analysis<sup>a</sup>:** |  |  |  |  |
| Any mRNA | 0.63[0.55 – 0.72] | 0.63[0.55 – 0.72] | 0.62[0.54 – 0.71] | 0.62[0.54 – 0.71] |
| BNT162b2 | 0.65[0.54 – 0.77] | 0.63[0.53 – 0.75] | 0.52[0.42 – 0.66] | 0.52[0.41 – 0.65] |
| mRNA-1273 | 0.60[0.49 – 0.75] | 0.62[0.50 – 0.77] | 0.72[0.60 – 0.85] | 0.72[0.60 – 0.85] |
| **Sensitivity analysis<sup>b</sup>** |  |  |  |  |
| Any mRNA | 1.00[0.80 – 1.25] | 1.01[0.80 – 1.27] | 1.10[0.82 – 1.47] | 1.10[0.82 – 1.47] |
| BNT162b2 | 1.01[0.75 – 1.37] | 1.00[0.74 – 1.35] | 0.93[0.61 – 1.43] | 0.91[0.59 – 1.39] |
| mRNA-1273 | 0.98[0.69 – 1.38] | 1.01[0.71 – 1.43] | 1.32[0.88 – 1.98] | 1.35[0.90 – 2.02] |

<sup>a</sup>Primary end point (significant health event): a new or worsening of a health event sufficient to cause work/school absenteeism, medical consultation and/or prevent daily activities in the previous seven days following each dose of vaccine
<sup>b</sup>Severe health event: a new or worsening of a health event resulting medical consultation in the previous seven days following each dose of vaccine
\*Reference: vaccinated non-pregnant women; adjusted for age groups and previous COVID-19 infection

